# SLO-MSNet: Discrimination of Multiple Sclerosis using Scanning Laser Ophthalmoscopy Images with Autoencoder-Based Feature Extraction

**DOI:** 10.1101/2023.09.03.23294985

**Authors:** Roya Arian, Ali Aghababaei, Asieh Soltanipour, Shwasa B Iyer, Fereshteh Ashtari, Hossein Rabbani, Raheleh Kafieh

## Abstract

**Background:** Optical coherence tomography (OCT) studies have revealed that compared to healthy control (HC) individuals, retinal nerve fiber, ganglionic cell, and inner plexiform layers become thinner in multiple sclerosis (MS) patients. To date, a number of machine learning (ML) studies have utilized Optical coherence tomography (OCT) data for classifying MS, leading to encouraging results. Scanning laser ophthalmoscopy (SLO) uses laser light to capture high-resolution fundus images, often performed in conjunction with OCT to lock B-scans at a fixed position, removing the effects of eye motion on image quality and allowing for evaluating the disease progression at follow-up examinations. To our knowledge, no ML work has taken advantage of SLO images for automated diagnosis of MS.

**Methods:** In this study, SLO images were utilized for the first time with the purpose of fully automated classification of MS and healthy control (HC) cases. First, a subject-wise k-fold cross-validation data splitting approach was followed to minimize the risk of model overestimation due to data leakage between train and validation datasets. Subsequently, we used several state-of-the-art convolutional neural networks (CNNs), including VGG-16, VGG-19, ResNet-50, and InceptionV3, as well as a custom CNN architecture trained from scratch. In the next step, we designed a convolutional autoencoder (CAE) to extract semantic features from the images which are then given as the input to four conventional ML classifiers, including support vector machine (SVM), k-nearest neighbor (K-NN), random forest (RF), and multi-layer perceptron (MLP).

**Results:** The custom CNN model outperformed state-of-the-art models with an accuracy (ACC) of 85%, sensitivity (SE) of 85%, specificity (SP) of 87%, and AUROC of 93%; however, utilizing a combination of the CAE and MPL yields even superior results achieving an ACC of 88%, SE of 86%, SP of 91%, and AUROC of 94%, while maintaining high per-class accuracies. The best performing model was also found to be generalizable to an external dataset from an independent source, achieving an ACC of 83%, SE of 87%, and SP of 79%.

**Conclusion:** For the first time, we utilized SLO images to differentiate between MS and HC eyes, with promising results achieved using combination of designed CAE and MLP which we named SLO-MSNet. Should the results of the SLO-MSNet be validated in future works with larger and more diverse datasets, SLO-based diagnosis of MS can be reliably integrated into routine clinical practice.

## 1. Introduction

Multiple sclerosis (MS) is an autoimmune disease of the central nervous system characterized by chronic inflammation, demyelination, and axonal degeneration (1). Currently, diagnosis of MS is on the basis of clinical presentations, magnetic resonance imaging (MRI) findings, and the presence of oligoclonal bands in the cerebrospinal fluid (CSF) (2). Notably, optical coherence tomography (OCT) studies have shown that peripapillary retinal nerve fiber layer (pRNFL) and both ganglionic cell and inner plexiform layers in the macular region (collectively abbreviated as mGCIPL) are thinner in MS patients compared with healthy controls (HCs) (3); a retrograde neuroaxonal atrophy following acute inflammatory attacks could be a likely explanation (4). The thickness of these layers has been associated with patients’ visual problems, MS subtypes, physical and cognitive disability, and MRI findings (5). Therefore, OCT parameters are now regarded as useful biomarkers for the quantitation of neurodegeneration in MS, allowing for a facilitated monitoring of disability progression and also assessing the efficacy of neuroprotective therapies (4). Artificial intelligence (AI), encompassing both machine learning (ML) and deep learning (DL), has emerged as a promising aid for diagnosing MS (6), with its impressive performance being shown in a recent meta-analysis (7). Data analyzed for the automated classification of MS primarily stem from MRI, serum, CSF, and OCT investigations (8); specifically, the OCT parameters have involved the macular and/or peripapillary thickness of RNFL, GCIPL, inner nuclear layer (INL), and the whole retina, alone or in combination (9–19), leading to high levels of ACC reaching up to 100% (14).

Infrared scanning laser ophthalmoscopy (SLO), also known as monochromatic fundus imaging, is another widely-used retinal imaging technology that uses low-powered laser light to create two-dimensional images of the retina. SLO is usually performed along with OCT B-scan acquisition; this allows for an accurate alignment of the B-scans despite eye movements, which improves the signal-to-noise ratio and reduces measurement variability at follow-up examinations (20) (Figure 1). Compared to fundus camera images that share a similar appearance, SLO images offer a higher resolution since the laser beam is passed through a confocal aperture and thus, light scattering is minimized (21), enabling a clearer and more detailed visualization of the retinal structures such as vessels and the optic nerve. Of note, when examining both fundus camera and SLO images, MS-related pathological changes have proven undetectable to human physicians, but could potentially be identified with the help of ML. Recently, ML models trained with ultra-wide (UWF) color SLO images have been remarkably successful in classifying several ophthalmological (22–31) and neurodegenerative diseases (32). Wisely et al. trained a Resnet50-based convolutional neural network (CNN) with retinal images captured via different modalities, including OCT, OCT angiography, and UWF color/fundus autofluorescence SLO images, finally achieving an AUC of 0.83. However, their model showed a poor performance when being fed solely with SLO images (32). Despite these advances, there remains a paucity of studies that assess the ACC of ML algorithms for automated diagnosis of neurodegenerative diseases, and in particular MS, using SLO images.

**Figure 1.**
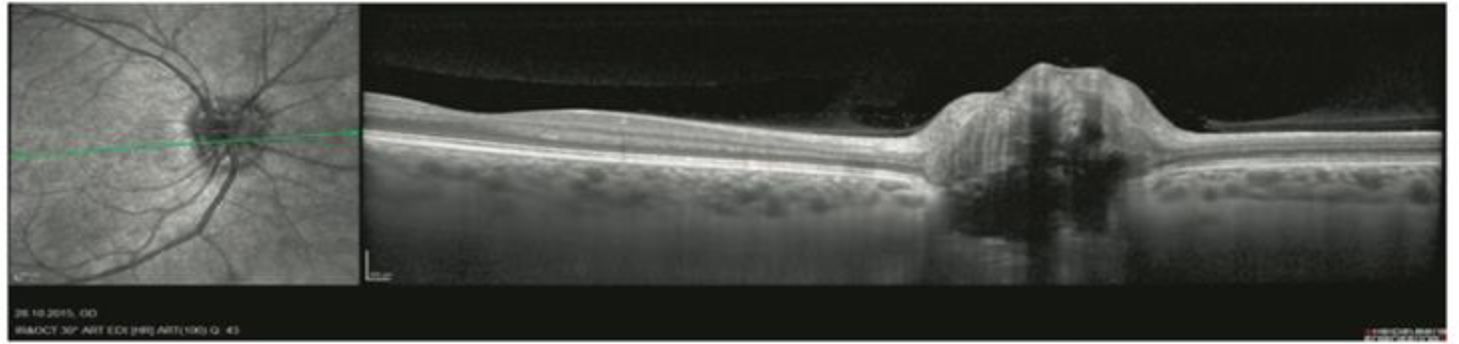
The acquisition window of the Spectralis software, containing both SLO image (left) and the OCT B-scan (right). Note that the green line superimposed on the SLO image corresponds to the position of the B-scan (reprinted from reference (33) under the terms of the Creative Commons Attribution 4.0 International License (34)).

To date, a number of studies have used OCT thickness measurements of different retinal layers for classification of MS using ML models (9–19). This is because a great body of evidence suggests that RNFL and GCIPL layers become thinner in MS patients, even in the absence of any history of ON (3–5). However, fundus images or other OCT data like B-scans have not been shown to be affected during MS course, at least in a way that can be noticed by human physicians. Of note, the current study is the first attempt to classify MS based on SLO images.

In the current study, we aimed to develop ML/DL models for classifying MS using infrared SLO images, obtained from a previous study including the OCT data of 53 MS patients and 85 HCs (35). To our knowledge, this would be the first attempt at SLO-based automated detection of MS. Moreover, our study would be among the few studies (14) that have used DL models for classifying MS based on retinal data.

The rest of this paper is arranged as follows: the “Methods and Materials” section (Section 2) describes the datasets, preprocessing steps, and the classification algorithms. The experimental results, including the appropriate plots, are reported in Section 3, with a discussion about them provided in Section 4. Finally, section 5 corresponds to the conclusion of the current work.

## 2. Methods and Materials

### 2.1. Dataset

The dataset utilized in this study consisted of two independent datasets, i.e., the Isfahan and Johns Hopkins datasets. Both datasets consisted of OCT scans and SLO images from patients with MS and HC individuals, captured using a SPECTRALIS® SD-OCT device (Heidelberg Engineering, Heidelberg, Germany). The incorporation of OCT technology with SLO in this device allows for locking the B-scans at a desired position using a real-time eye tracking system (TruTrack™), which removes the effects of eye motion during image acquisition, creating high-quality OCT images. Furthermore, this is necessary for an accurate evaluation of the disease progression since a B-scan on the same position should be studied during follow-up examinations (20). The Isfahan dataset was obtained from a previous study by Ashtari et al. (35), consisting of a total of 314 SLO images from 53 patients with MS (164 images) and 85 HC individuals (150 images). The study was undergone between April 2017 and March 2019 at Kashani Comprehensive MS Center, Isfahan, Iran, a main referral center for MS in Isfahan. The Johns Hopkins dataset was publicly available, consisting of SLO and OCT images of the right eyes from 35 individuals, 14 HCs and 21 patients with MS (36), with a different demographic and clinical background compared to those in the Isfahan dataset.

#### 2.1.1. Train and Test Splitting

For the Isfahan dataset, random train and validation data splitting was performed using k-fold cross-validation (CV). K-fold CV is preferred to random split in terms of completeness and generalization, since it ensures that the system has seen the complete dataset for training and it is guaranteed to both train and test sets on every observation of the dataset an equal number of times (k−1 and 1 times, respectively), while in random split by re-sampling at each iteration, duplicate members of the test set can be selected twice, or even more. K-fold CV is a more preferable choice over random data splitting because it guarantees that the entire dataset is used for training the model, and each observation appears an equal number of times, i.e., k-1 times in training and 1 time in testing phases. On the other hand, random split may lead to duplicate selections in the test set due to re-sampling at each iteration.

Stratified sampling was also used to ensure that each fold has the same proportion of samples with a certain label, i.e., MS or HC. Furthermore, to prevent data leakage between train and validation datasets, a “subject-wise” approach was followed that involves putting all images belonging to each individual, regardless of its left-or-right orientation, in a single group. Therefore, the images of the same participant cannot be used in both training and validation datasets concurrently, preventing an overestimation of the model performance (37).

#### 2.1.2. Data Augmentation

Data augmentation is a popular preprocessing technique in ML studies with limited training data in order to minimize the risk of overfitting. It works by adding minor modifications to the original input images to create new but similar examples, artificially increasing the size and diversity of training samples. In this study, several geometric and color space transformations were applied to the SLO images, which consisted of vertical flip, height shift in the range of ±5 pixels, width shift in the range of ±30 pixels, rotation in the range of ±5 degree, zoom in range of ±0.2 and image brightness in the range of 0.2 to 1.5.

### 2.2. Classification

Of note, CNNs work by integrating the dimensionality reduction and classification processes in an end-to-end manner. Using a CNN, features are extracted from input images through the convolutional layers, with no external feature extraction method needed to be applied; the resulting features are then utilized for final classification made by the FC layers (45). To test whether even higher classification accuracies can be achieved, an independent feature extraction method that is suitable for image data (CAE) was applied on the SLO dataset and the obtained features were classified with ML algorithms.

#### 2.2.1. Deep Learning (CNNs)

In this study, we took advantage of transfer learning by using a number of state-of-the-art CNN architectures, namely VGG-16 (38), VGG-19 (38), ResNet-50 (39), and InceptionV3 (40), which have yielded high levels of classification accuracies on large image benchmarks like ImageNet (41). The idea behind transfer learning is that instead of learning from scratch, the knowledge learned by such high-performing architectures can be transferred to a new dataset with a much lower size, thereby preventing overfitting. In this study, the above architectures were utilized as a fixed feature extractor, where the weights of the models were frozen; however, the fully connected (FC) part was replaced by a custom one applicable to our binary classification task (MS vs HC). Also, the fine-tuning approach was applied to the CNN model with the best performance; the topmost convolutional layers that capture features specific to the new domain became unfrozen so that the weights of these layers can be updated during the training process. However, the first convolutional layers were still kept frozen since they detect general features shared between natural images found in the ImageNet dataset and SLO images. Furthermore, a custom CNN model was developed to be fully trained on our dataset.

In order to search for the optimal CNN hyperparameters, including learning rate, batch size, dropout probability, number of hidden layers of the FC part, and number of neurons in each hidden layer, the Optuna hyperparameter optimization framework was utilized (42); the final architecture is shown in Figure 2. A brief introduction of CNNs is provided in the Supplementary Material on pages 2 and 3.

**Figure 2.**
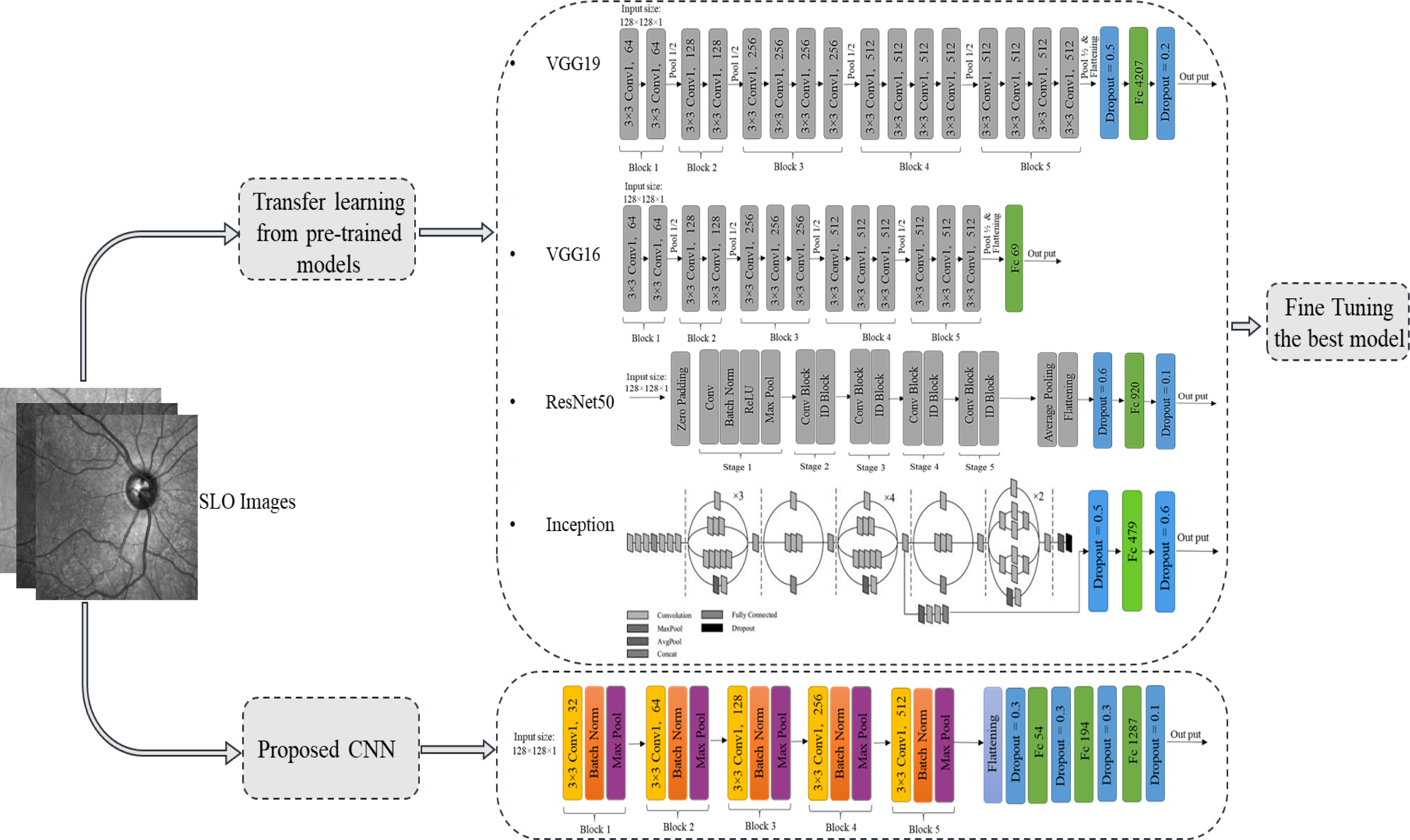
Overview of MS classification based on scanning laser ophthalmoscopy (SLO) images using convolutional neural networks (CNNs). The gray blocks represent untrainable layers with frozen weights and the colored blocks show trainable layers.

As shown in Figure 2, the proposed CNN compromised five blocks, each containing 2D convolutional layer (with a kernel size of 3 × 3), ReLU activation function, batch normalization, and max pooling layers stacked together; the number of channels (feature maps) in each convolutional layer progressed from 32, 64, 128, 256, and finally 512 through the network. Finally, an FC part consisting of three hidden layers with 54, 194, and 1287 neurons was added to the end of the CNN, with ReLU and sigmoid as the activation functions for the first three layers and the output layer, respectively. To reduce the risk of overfitting, dropout regularization with a probability of 0.3 was applied just after flattening layer were given to the FC part and to the first two FC layers as well. A dropout rate of 0.1 was also added at the end, just before the output layer (Figure 2). Adam and binary cross entropy loss were used as the optimizer and loss function, respectively.

#### 2.2.2. Feature Extraction and Machine Learning Classifiers

Dimensionality reduction is a necessary preprocessing step when dealing with high-dimensional data like images in order to overcome the so-called “curse of dimensionality”, a problem leading to high computational costs and poor performance of ML models. To address this, a number of methods have been developed that project data points to a lower dimensional space which still retains the most important information of the original data, e.g., principal component analysis (PCA) (43) and autoencoder neural networks (AEs) (44); the resulting features can then be given to ML algorithms for final classification. Indeed, when working with limited data, utilizing a combination of feature extraction and conventional machine learning algorithms is expected to outperform deep neural networks.

AEs aim to generate a compressed and meaningful representation of input data by learning to reconstruct it in an unsupervised manner. AEs consist of an encoder and a decoder, which are neural networks (NNs) in most scenarios, and a bottleneck layer between the two. The encoder network tries to map input data to a latent feature space with a lower dimensionality (bottleneck layer) and the decoder will then utilize the resulting features to create output data as much similar to the original data. During training, AEs minimize the difference between the original and reconstructed data, often using mean square error as the loss function for such a regression problem. Some of the various applications of AEs include image denoising, image generation, anomaly detection, and feature extraction; the latter can be seen as a non-linear extension of PCA. Indeed, in cases where no non-linear activation functions are used in AEs, the latent feature space created by the encoder is the same as the PCA output (44).

Interestingly, combining AEs with CNNs seems to yield more informative representations for image data by preserving the two-dimensional features of images, e.g., edges and corners. In the current study, the resulting features from such CAE were then given to a SVM classifier. Subsequently, CAE hyperparameters like the number of convolutional and pooling layers, number of filters, kernel size, learning rate, batch size, and dropout probability were optimized empirically to get the highest possible classification accuracies (without considering the reconstruction error). Furthermore, the features obtained from the CAE were also used to train a number of other ML classifiers, namely, K-NN, MLP, and RF to achieve the highest possible level of accuracy. Each of these three classifiers is briefly introduced in the Supplementary Material on page 1. In this study, the grid search algorithm and the Optuna library were utilized to find the optimal hyperparameters for each classifier. Grid search algorithm involves calculating the ACC of each combination of all specified hyperparameters and then selecting the best value for them (45) while Optuna uses Bayesian optimization to find the optimal set of hyperparameters. Figure 3 provides an overview of this section (feature extraction plus ML classification), including the architecture of the designed CAE. All Conv2D layers in the CAE employed the ReLU activation function, except for the final layer where a sigmoid function was applied. Additionally, the optimization process was carried out using the Adam optimizer, and the loss was computed using the Huber loss function. The mean squared error (MSE) is a great loss function for learning outliers while the mean absolute error (MAE) ignores them. However, the Huber loss function demonstrates lower sensitivity to the outliers compared to the MSE by balancing the MSE and the MAE together. It behaves quadratically (like MSE) for small values of the difference between the predicted value *y* and the actual value *f(x)*, and linearly (like MAE) for large values (46). The mathematical formula for Huber loss function is as follows:

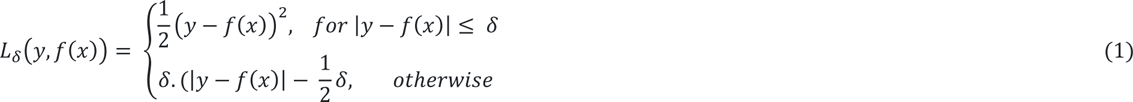

**Figure 3.**
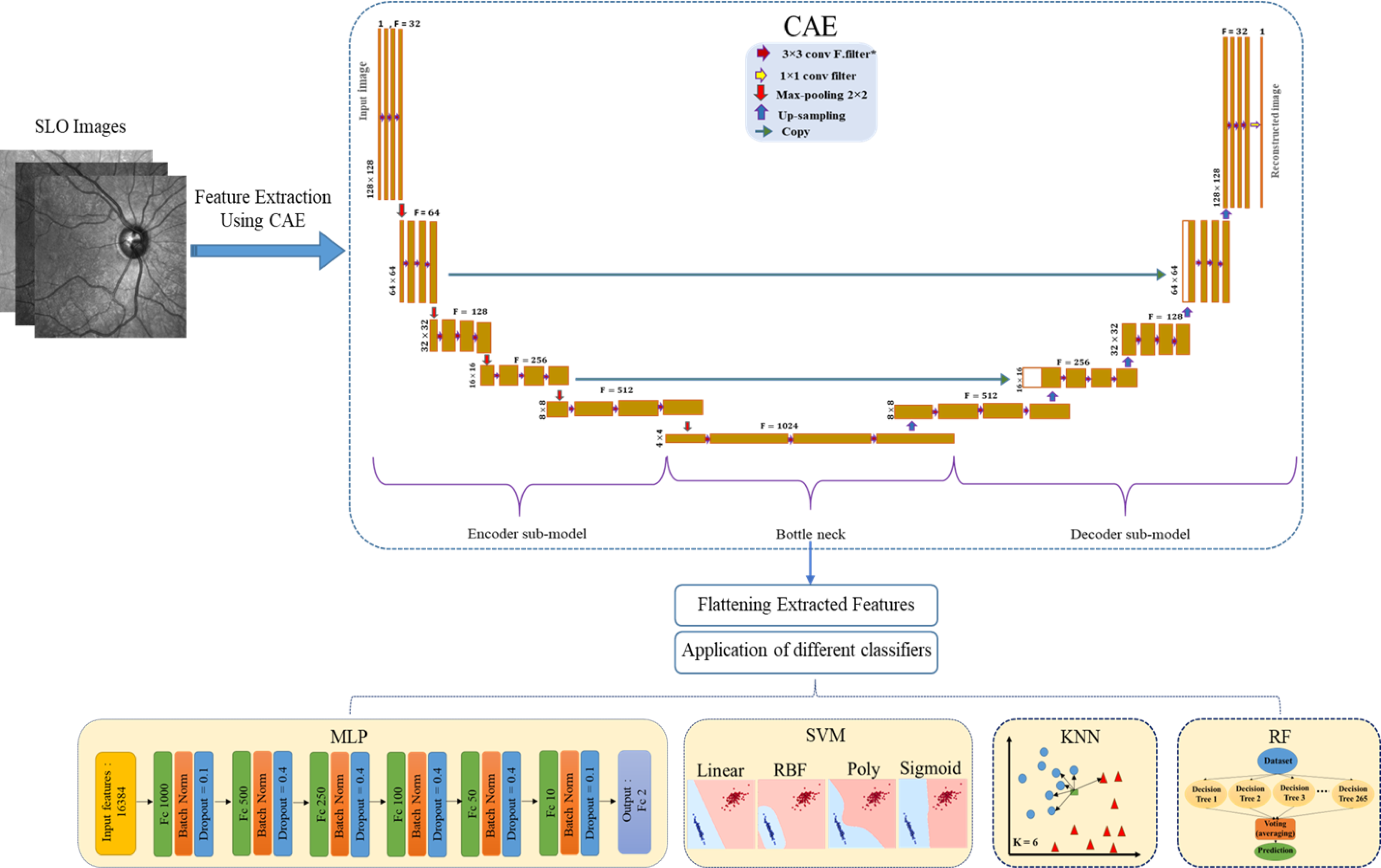
Overview of MS classification based on SLO images using a CAE as the feature extractor followed by conventional machine learning classifiers; MLP, multi-layer perceptron; SVM, support vector machine; K-NN, k-nearest neighbor; RF, random forest

Where the hyperparameter *δ* introduces a threshold which was set to 1 in this study.

### 2.3. Evaluation of Classification Models

The metrics that were employed to evaluate the performance of each classifier consisted of ACC, SE, SP, precision (PR), and F1-score, with the mathematical formula for calculating them being represented as follows:

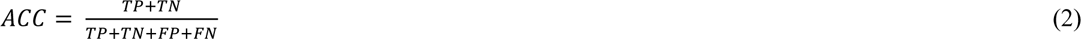

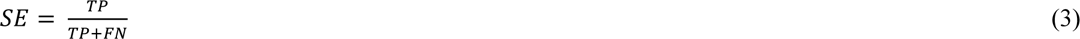

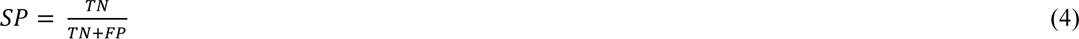

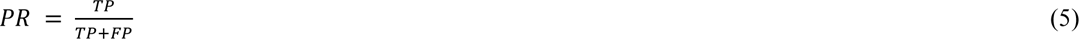

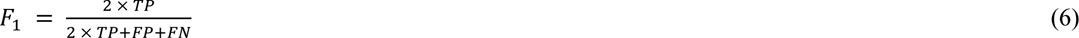

Where TP, FN, TN, and FP are the abbreviated forms of true positive, false negative, true negative, and false positive, respectively. Moreover, the receiver operating characteristics (ROC) and precision-recall curves were plotted; the ROC curve illustrates the relationship between the true positive and false positive rates, while the precision-recall curve showcases the tradeoff between the precision and recall across various thresholds. The areas under these curves, known as area under the ROC curve (AUROC) and precision-recall area under the curve (AUPRC), were also calculated Moreover, the gradient-based class activation map (Grad-CAM) (47) and the gradient-weighted latent activation mapping (Grad-LAM) (48), i.e., an extension of the Grad-CAM that was introduced for unsupervised representation learning, were utilized to obtain saliency heat maps for interpreting CNN and CAE predictions, respectively.

## 3. Results

All the experiments in this study were implemented using Python programming language, in the Keras platform backend in python 3.7 software environment.

Overall, 314 SLO images from the Isfahan dataset, consisting of 85 and 53 “subject” groups for HC and MS individuals, respectively, were utilized to train CNN (Section 3.1.) and CAE plus ML classifiers (Section 3.2.). All images were resized to have a 128 × 128 × 1 pixel size, and the pixel intensities were divided by 255 to have values ranging from 0 to 1. Furthermore, all images belonging to the left eyes were mirrored.

### 3.1. Deep Learning (CNNs)

As mentioned in Section 2.2.1, state-of-the-art CNN models were first used as feature extractors with the weights of all layers kept frozen, followed by FC layers with a custom number of hidden layers containing varied number of neurons (Figure 2). VGG-19 (with 14,714,688 untrainable parameters) showed the best performance, with a mean ACC of 84% (SE = 83%, SP = 87%, PR = 87%, F1-score = 84%, AUROC = 92%) over five consecutive executions (Table 1). Moreover, the custom CNN model that was trained from scratch showed more desirable results (ACC = 85%, SE = 85%, SP = 87%) (Table 1).

**Table 1.** Performance metrics of the state-of-the-art and the proposed convolutional neural networks (CNNs) for classification of MS using SLO images. Best results are bolded, revealing that the custom CNN is the winning classifier.

| Model | ACC | SP | SE | PR | F1 | AUROC | AUPRC | Normal class ACC | MS class ACC | Optimal hyper parameters |  |
| --- | --- | --- | --- | --- | --- | --- | --- | --- | --- | --- | --- |
|  |  |  |  |  |  |  |  |  |  | Batch size | Learning rate |
| **InceptionV3** | 0.81 | 0.83 | 0.80 | 0.83 | 0.81 | 0.90 | 0.92 | 0.83 | 0.80 | 16 | 6e-4 |
| **ResNet-50** | 0.71 | 0.84 | 0.71 | 0.83 | 0.70 | 0.79 | 0.85 | 0.84 | 0.70 | 64 | 1.4e-4 |
| **VGG16** | 0.83 | 0.86 | 0.81 | 0.85 | 0.83 | 0.92 | 0.93 | 0.86 | 0.81 | 8 | 1.7e-4 |
| **VGG19** | 0.84 | **0.87** | 0.83 | **0.87** | 0.84 | 0.92 | **0.94** | **0.87** | 0.83 | 8 | 1.2e-4 |
| **Proposed CNN** | **0.85** | **0.87** | **0.85** | **0.87** | **0.85** | **0.93** | **0.94** | **0.87** | **0.85** | 16 | 2.5e-4 |

To further improve the results, we also unfreeze the first, second, and fourth topmost convolutional layers of VGG19 (as the winning transfer learning algorithm), so 2,359,808, 4,719,616, and 7,079,424 parameters can be re-trained with the SLO image dataset, respectively. However, this fine-tuning approach did not lead to a better performance. For instance, when the first topmost layer was unfrozen, the obtained results were as follows: ACC = 84%, SE = 83%, SP = 85%, PR = 85%, AUROC = 92%, F1-score = 83%) (Figure A in the Supplementary Material).

Box plots for the state-of-the-art and the custom CNN models are drawn in Figure 4.

**Figure 4.**
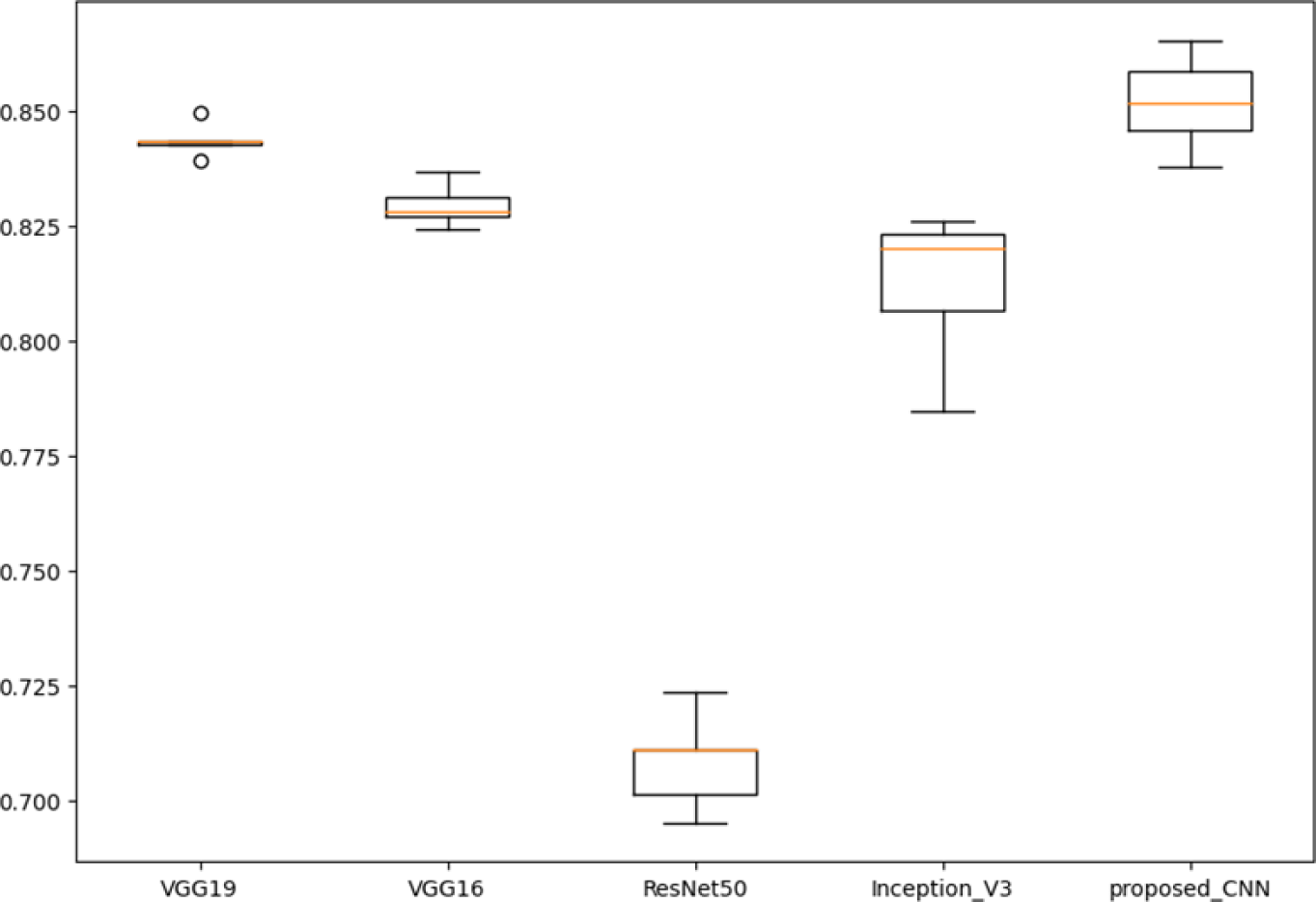
Box plots for state-of-the-art convolutional neural networks (CNNs) and the proposed model, drawn following five repeated executions.

### 3.2. Feature extraction and Machine Leaning Classifiers

The proposed architecture for the CAE is illustrated in Figure 3, with a brief explanation available in the Supplementary Material on page 3. The flattened representation of the bottleneck output, suggestive of the most important features of SLO images, was then utilized for classifying MS and HC using SVM, MLP, RF, and K-NN algorithms. The SVM classifier achieved an ACC of 86% (SE = 82%, SP = 90%) using a radial basis function (RBF) kernel as the winning kernel (Table A in the Supplementary Material); the optimal values for CAE hyperparameters, including batch size (= 16), learning rate (= 1e-5), and dropout probability (= 0.2), were then kept unchanged when other classifiers were utilized. Furthermore, to visualize which regions have the most contribution to the CAE reconstruction score, the saliency maps of several MS and HC images from the test dataset were created using Grad-LAM, illustrated in Figure 5. Comparison of These heat maps and the heat maps obtained by the custom CNN (using the Grad-CAM algorithm) for the same MS and HC images can provide some perspective of the regions of the images with most impact for each model (Figure 5).

**Figure 5.**
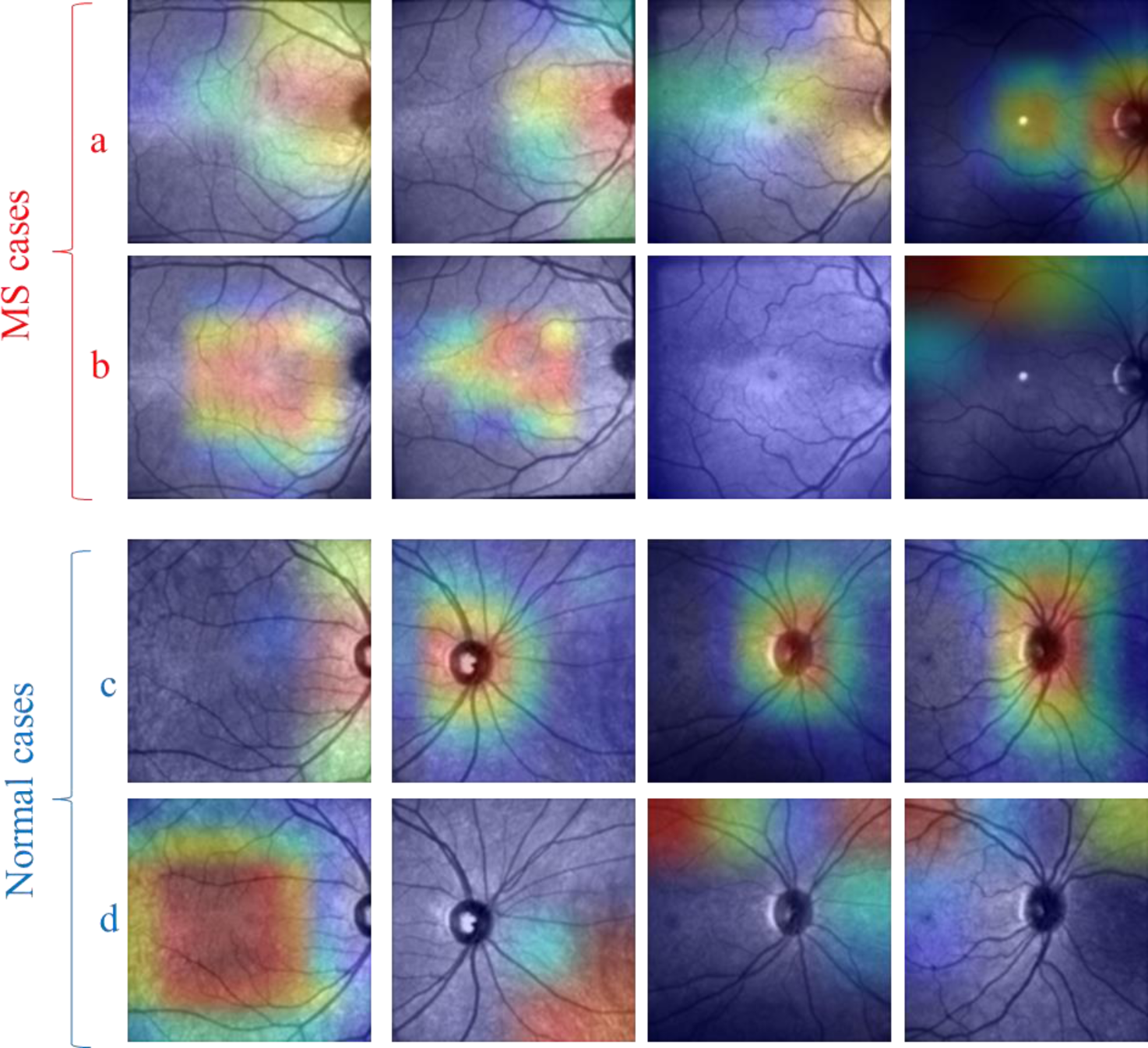
Saliency maps generated for four SLO images of MS and HC subjects from the test dataset when the proposed CAE (a and c) and CNN (b and d) were utilized.

Subsequently, other conventional ML algorithms, namely, K-NN, MLP, and RF, were applied to the CAE-extracted features, with MLP achieving the best results (ACC= 88%, SE= 86%, SP= 91%; PR= 90%, F1-score= 88%; AUROC = 94%; AUPRC = 95%) (Table 2). Figure 6 shows the ROC/precision-recall curves and the confusion matrices for all the four classifiers. The best hyperparameters of K-NN, MLP, and RF are depicted in the Supplementary Material Table B. The high discriminant capacity of the features extracted by the CAE is visualized in Figure 7 using Uniform Manifold Approximation and Projection (UMAP) for dimensionality reduction technique (49) In the same figure, the discriminant capacity of the CAE is also compared with that of PCA as a simple feature extraction technique where the number of components was set to 300.

**Figure 6.**
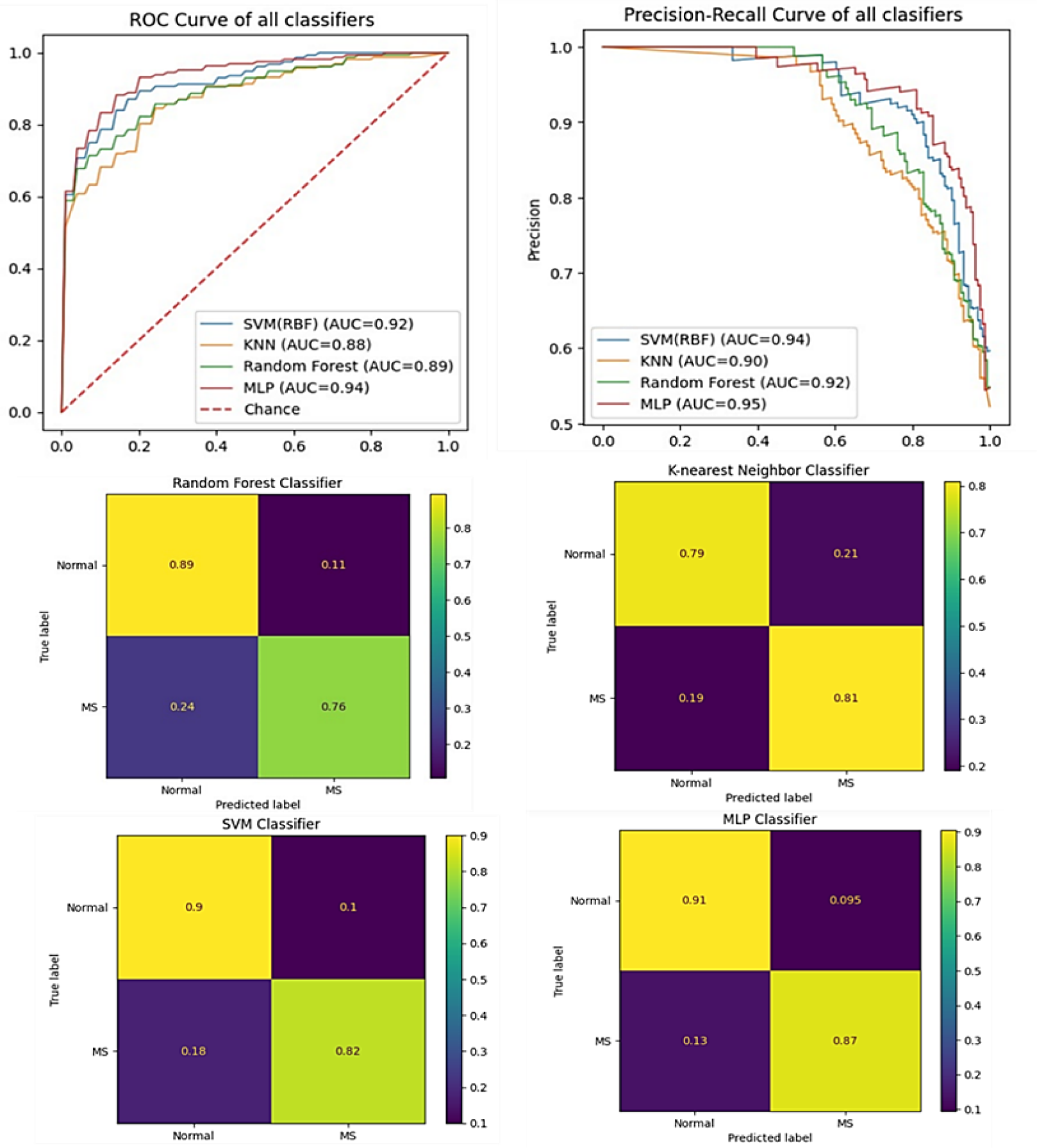
Receiver operating characteristics (ROC) curve (top left), precision-recall curve (top right), and the confusion matrices (bottom) for the four machine learning classifiers. MLP, multi-layer perceptron; SVM, support vector machine; K-NN, k-nearest neighbor; RF, random forest

**Figure 7.**
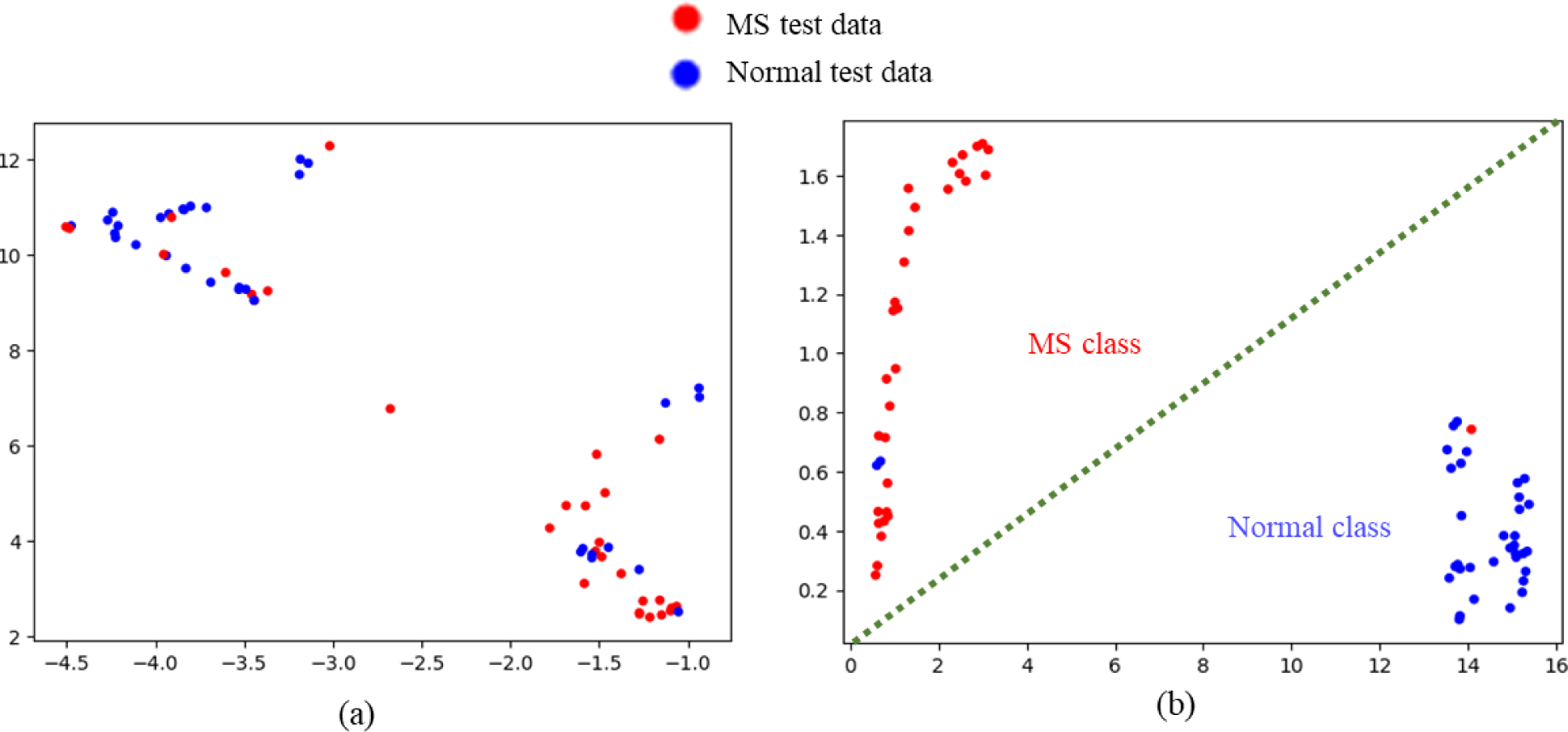
Visualization of the features obtained from the convolutional autoencoder neural network using Uniform Manifold Approximation and Projection (UMAP) for dimensionality reduction technique (49)

**Table 2.** Performance metrics of the four machine learning models for classifying MS using scanning laser ophthalmoscopy (SLO) images. The classifiers were applied on the features extracted using the proposed convolutional autoencoder, with MLP being the winning model.

| model | ACC | SE | SP | PR | F1 | AUROC | AUPRC |
| --- | --- | --- | --- | --- | --- | --- | --- |
| RF | 0.83 | 0.76 | 0.89 | 0.92 | 0.82 | 0.9 | 0.92 |
| K-NN | 0.8 | 0.81 | 0.79 | 0.8 | 0.8 | 0.88 | 0.91 |
| RBF-SVM | 0.86 | 0.82 | 0.9 | 0.89 | 0.86 | 0.92 | 0.94 |
| **MLP** | **0.88** | **0.86** | **0.91** | **0.9** | **0.88** | **0.94** | **0.95** |

The classification ACC of these ML models for each class individually was also calculated and compared with that of CNNs, with the corresponding bar charts plotted in Figure 8.

**Figure 8.**
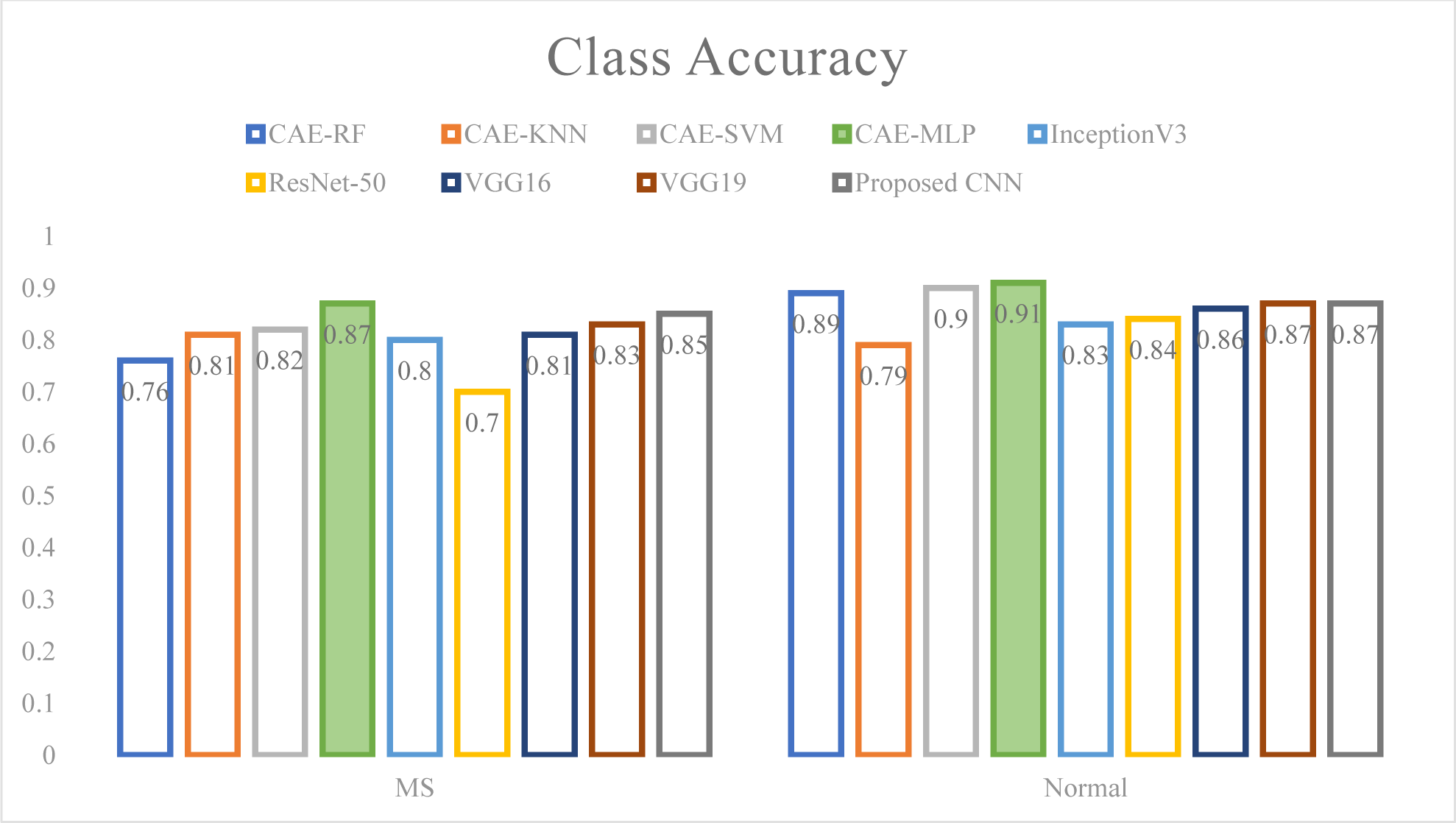
Classification ACC of MS and HC classes individually using CNNs and combination of CAE and ML classifiers.

In addition to the subject-wise approach mentioned in Section 2.1.1, two other data splitting methods were also applied, namely record-wise and eye-wise approaches. The record-wise approach is a conventional data-splitting method, in which the entire image dataset (314 images) was given to the 5-fold CV algorithm to create training (4/5 × 314 ≈ 251) and validation (1/5 × 314 ≈ 63) images in each fold. In the eye-wise approach, all images belonging to either the right or left eye of each individual were put in separate groups; 150 and 88 “eye” groups were resulted for HC individuals and patients with MS, respectively. Results of three approaches are summarized in Table 3.

**Table 3.** performance metrics of the support vector machine (SVM) with different kernels when applied to features extracted by the convolutional autoencoder neural network using three train/validation data splitting approaches. Best results are bolded.

| Data splitting approach | SVM kernels | ACC | SE | SP | PR | F<sub>1</sub> | AUROC | AUPRC |
| --- | --- | --- | --- | --- | --- | --- | --- | --- |
| Record-wise | Linear | 0.89 | **0.97** | 0.80 | 0.83 | 0.88 | 0.98 | 0.98 |
|  | Polynomial | 0.84 | 0.78 | **0.90** | **0.89** | 0.84 | 0.94 | 0.95 |
|  | RBF | **0.91** | **0.97** | 0.83 | 0.85 | **0.91** | **0.98** | **0.99** |
|  | Sigmoid | 0.65 | 0.78 | 0.50 | 0.61 | 0.64 | 0.77 | 0.81 |
| Eye-wise | Linear | 0.87 | **0.89** | 0.87 | 0.86 | 0.87 | 0.93 | **0.95** |
|  | Polynomial | 0.84 | 0.84 | 0.83 | 0.83 | 0.84 | 0.90 | 0.93 |
|  | RBF | **0.89** | 0.85 | **0.92** | **0.92** | **0.88** | **0.95** | **0.95** |
|  | Sigmoid | 0.70 | 0.76 | 0.60 | 0.65 | 0.70 | 0.80 | 0.85 |
| Subject-wise | Linear | 0.85 | **0.86** | 0.84 | 0.85 | 0.85 | **0.93** | 0.93 |
|  | Polynomial | 0.84 | 0.83 | 0.85 | 0.85 | 0.84 | 0.92 | 0.93 |
|  | RBF | **0.86** | 0.82 | **0.90** | **0.90** | **0.86** | **0.93** | **0.94** |
|  | Sigmoid | 0.71 | 0.61 | 0.83 | 0.79 | 0.71 | 0.78 | 0.84 |

### 3.3. Ablation Study

It is worth mentioning that an ablation study, investigating the effect of feature extraction prior to the classification, was also undertaken when SVM with an RBF kernel and MLP were utilized. The SVM classifier (with RBF kernel) separate data points by projecting them to an infinite dimensional kernel space(50) ; thus, it may not hypothetically seem reasonable to reduce the dimensionality of our dataset before applying this algorithm. Also, NNs generally exhibit superior performance when dealing with higher dimensional data, not passed through feature extraction/selection algorithms (51). Therefore, classification performance of SVM (with RBF kernel) and MLP was evaluated with and without applying the dimensionality reduction method. Results are summarized in Table 4.

**Table 4.** results of the ablation study undertaken MLP and the SVM with radial basis function kernel (RBF-SVM) with and without feature extraction prior to classification. Best results for each classifier are bolded. Using CAE in both methods yields higher results.

| Model | CAE | ACC | SE | SP | PR | F<sub>1</sub> | AUROC | AUPRC |
| --- | --- | --- | --- | --- | --- | --- | --- | --- |
| MLP | Used | **0.88** | **0.86** | **0.91** | **0.9** | **0.88** | **0.94** | **0.95** |
|  | Removed | 0.79 | 0.78 | 0.82 | 0.82 | 0.79 | 0.88 | 0.9 |
| RBF-SVM | Used | **0.86** | **0.82** | **0.9** | **0.89** | **0.86** | **0.92** | **0.94** |
|  | Removed | 0.81 | 0.82 | 0.81 | 0.81 | 0.81 | 0.88 | 0.9 |

### 3.4. Generalization ability on the external dataset

To check for the generalization ability of the optimal model, i.e., CAE plus MLP classifier, we used the Johns Hopkins dataset as an extra source for the test phase. Our results were shown to be promising (ACC = 83.1%, SE = 86.5%, SP = 79.4%, PR = 80.1%, F1-score = 83.1%, AUROC = 93.1%, AUPRC = 94.1%).

## 4. Discussion

According to the Table 1, the custom CNN model that was trained from scratch showed more desirable results (ACC = 85%, SE = 85%, SP = 87%). This could partly be attributed to the fact that the architecture of our proposed CNN was not as complex compared to that of the state-of-the-art models, which is probably more suitable for the SLO images containing relatively simple patterns to be recognized. For the same reasons, the fine-tunning approach did not lead to a superior performance compared to the transfer learning given an extreme increase in the number of trainable parameters.

Applying a CAE for feature extraction showed a superior performance compared to CNNs; therefore, we speculate that to reconstruct the input image effectively, the CAE should learn more semantically meaningful representations. The idea behind this hypothesis is similar to the paper published by Pathak et al (52), introducing the “context encoders” as a solution for semantic inpainting. Context encoders are indeed a type of CAEs that are aimed to generate missing parts of an image with respect to the contextual information from the surrounding regions. The latent layer of a context encoder was shown to provide valuable features that serve as reliable indicators of the input images, resulting in appealing results across different tasks, including object detection, semantic segmentation, and classification (52). Furthermore, the capability of the proposed CAE in detecting more informative representations could be attributed, at least partly, to the connections between two of the encoder and decoder blocks. This will propagate the information between encoder and decoder parts and prevent the precise information to be lost during the up-sampling process, since a feature map with higher resolution is constructed and then processed by the decoder convolutional layers. In addition, as illustrated in Figure 5, the optic nerve head and the area around it in MS undergoes pathological changes that can be detected by the proposed CAE; however, these pathologies remain unnoticed by human physicians. Of note, the custom CNN trained from scratch, identified other regions than those adjacent to the optic nerve captured by the CAE.

According to Figure 8, the proposed CAE feature extraction model combined with the MLP classifier was revealed to outperform other methods in classifying both MS and HC classes individually and provide a higher per-class accuracy.

According to the Table 3, as expected, the best results were achieved using the record-wise approach, followed by eye-wise and subject-wise data splitting methods. In both record-wise and eye-wise approaches, it is possible to simultaneously utilize image (s) of a participant as the training data and the other image (s) belonging to the same person as the validation data; therefore, the subject-wise method seems to be a more reliable data-splitting approach. This is because this strategy is potentially protected against an overestimation of the model performance due to the absence of data leakage between train and validation datasets (37). As shown in the Supplementary Material Table D, should a conventional record-wise approach be employed, a best ACC of 91% would be achieved, which is comparable to prior OCT studies with most of them using the same technique for train and validation data-splitting. The only exception is the study by Khodabandeh et al. (18) who used the Isfahan dataset, similar to the current study, which allow us to directly compare the results of these two studies; however, Khodabandeh et al. retrieved the OCT scans instead of SLO images. They cropped 20 × 20, 30 × 30, and 40 × 40 squares around the macula from the thickness maps of different retinal layers and applied PCA and recursive feature elimination for dimensionality reduction; finally, three conventional ML classifiers, namely SVM, RF, and MLP were trained with the obtained features. The authors were able to reach an ACC of 88% using an SVM with a linear kernel that was applied to the PCA-extracted features from the GCIPL/INL thickness maps. The best performing model in the current study also yielded a similar accuracy, suggesting that utilizing SLO instead of OCT could be a promising area of research for automated classification of MS.

According to Table 4, Results of ablation study revealed that by removing the feature extraction step, the classification ACC dropped from 86% to 81% and 88% to 79% for SVM and MLP classifiers, respectively. Therefore, feature extraction seems to be an essential preprocessing step in for the studied data.

As reported in Section 3.4, the model proposed in this study can reliably be applied on the SLO data from independent centers including individuals of other ethnicities with different demographic and clinical profile Two other multi-center studies that used OCT thickness data for classifying MS have been performed by Kenny et al. (19) and Khodabandeh et al. (18). The test dataset in the latter study was also desirably classified using the model trained with their first dataset (Isfahan dataset).

During recent years, automated diagnosis of MS has been made possible using ML algorithms, with remarkable overall ACC of 94% (7). Various input data have been utilized thus far, with the most desirable results achieved using the parameters obtained from MRI (pooled ACC = 96%), OCT (pooled ACC = 93%), CSF/serum (pooled ACC = 93%), and even gait and breathing pattern (pooled ACC = 88%) investigations. The OCT-based studies have mainly applied conventional ML classifiers on the thickness values (9,10,12,13,15–17,19) or the extracted features from them (11,14,18), with only one study utilizing DL (14). López-Dorado et al. employed Cohen’s d coefficient technique on the thickness maps of RNFL, GCL+ (equivalent to GCIPL), GCL++ (equivalent to GCIPL plus RNFL), the total retina, and the choroid from 48 MS patients and 48 HC individuals. The resulting thickness map images were then given to a custom CNN model made up of two successive blocks each containing one convolutional and one pooling layer, ultimately achieving very encouraging results (SE = 100%, SP = 100%). Similar to López-Dorado et al.’s, three other studies captured OCT data using a swept-source device (SS-OCT) (9,10,12), all leading to ACC levels of more than 90%. Such promising results could partly be attributed to the high-resolution scans generated by the SS-OCT technology. However, it should be noted that these studies had a limited sample size with insufficient diversity; indeed, three studies used the same dataset (9,12,14). The largest study that aimed to classify MS based on OCT data was undergone by Kenny et al. (19), consisting of 1568 MS patients and 552 HC subjects from the United States, Europe, and the Middle East. The dataset included various demographic, visual acuity, and SD-OCT parameters; using classification and regression tree models, the authors identified GCIPL thickness of both eyes in average, inter-eye GCIPL thickness difference, and binocular 2.5% low-contrast letter acuity as the features with the highest discriminant capacity. Kenny et al. applied both logistic regression and SVM algorithms that ultimately were shown to have a similar performance. Using SVM with a linear kernel, an ACC of 88%, a SE of 83%, and a SP of 90% were achieved. Overall, although the majority of ML research on MS classification has taken advantage of MRI (8), OCT measurements have also been shown to be invaluable input data. Notably, the diagnostic performance of the models trained with MRI and OCT parameters are not far different, but the OCT technology is much less invasive and costly. In the current study, we utilized SLO images instead of OCT data, resulting in a best ACC of 88%. As mentioned above, the proposed model, which we name SLO-MSNet, is indeed a combination of CAE for feature extraction and a MLP for final classification.

This study had several limitations. Although the generalizability of the proposed model was confirmed using an independent dataset, the train and validation images were from a single center and consisted of a limited number of samples. Also, we were not able to separate the eyes with a prior history of ON which are shown to have a thinner RNFL and GCIPL compared to the ON-negative eyes. Hypothetically, SLO images of the ON-positive eyes might also become more affected and needed to be separated to have more robust results. Furthermore, SLO images could be incorporated with fundus images and OCT data such as thickness measurements, thickness map images, or even projection images. Projection images are typically generated by taking the average of OCT A-scan intensities between the retinal layer boundaries (53). With this increased variety of information given to the model, more favorable outcomes could potentially be attained. Finally, the cross-sectional nature of this study precludes any conclusion regarding the disease progression.

## 5. Conclusion

To conclude, we have taken a significant step towards automated and precise detection of MS using a non-invasive, low-cost, and easily accessible technology like SLO. This is of great importance since in current clinical practice, diagnosis of MS is a challenging and time-consuming task, heavily relying on the findings from MRI and CSF investigations. We observed that the proposed CAE with connections between some of the encoder and the decoder paths outperforms state-of-the-art CNNs, probably because it yields more informative feature representations. In order to enhance the reliability and real-world applicability of our findings, we employed a subject-wise train and validation data-splitting strategy during K-fold cross-validation. Additionally, we assessed the performance of the SLO-MSNet on an external dataset obtained from a different source to test its generalizability. Indeed, future multi-center studies are encouraged to further evaluate the diagnostic accuracy of ML algorithms trained with SLO images, paving the way for their integration into routine clinical practice.

## Data and code Availability

Code and models are available at: https://doi.org/10.5281/zenodo.8217281

Optuna code to find optimal hyperparameters is also available at: https://doi.org/10.5281/zenodo.8218403

## Data Availability

the data is not available. However, codes are available at https://doi.org/10.5281/zenodo.8217281 and https://doi.org/10.5281/zenodo.8218403.

https://doi.org/10.5281/zenodo.8218403

