## Supplementary for "SLO-MSNet: Discrimination of Multiple Sclerosis using Scanning Laser Ophthalmoscopy Images with Autoencoder-Based Feature Extraction"

**Title Page**

#### A brief introduction of the machine learning classifiers that were applied to the extracted features from the SLO images

##### Support vector machine

Support vector machine (SVM) is a widely used supervised learning algorithm in classification and regression problems. It works by finding an optimal hyperplane that maximizes the decision boundary between different data classes. For data points that are not linearly separable, kernel functions, such as radial basis function (RBF), sigmoid, and polynomial, are utilized, increasing the dimensionality of feature space (1).

##### K-nearest neighbor

k-nearest neighbor (K-NN) is a simple and easily understandable ML algorithm that aims to classify new data based on the classes of its nearest training examples. Essentially, K-NN identifies the k closest neighboring examples to the input data with an unknown class and assigns a label based on the most common class among those nearby samples. Since it requires the training examples to be available during runtime, K-NN is considered a lazy learning technique. To optimize its performance, two hyperparameters needed to be set for K-NN: the number of nearest neighbors (k) and the choice of the distance metric that is utilized to determine similarity (2).

##### Multilayer Perceptron

Multilayer perceptron (MLP) is a traditional neural network, which consists of several interconnected processing units, also called neurons that are organized in consecutive layers including an input layer, one or more hidden layers, and ultimately, an output layer. All nodes in hidden layers utilize a nonlinear activation function. If the output of a neuron exceeds a threshold, that neuron becomes activated and sends signals to the neurons of the next layer, similar to the way biological neural cells are connected together. Training of an NN involves updating the weights between the neurons in a way that the final prediction made by the output layer becomes closer to the actual class of the input data, through a process known as backpropagation The number of hidden layers, the number of neurons in each layer, and the activation functions used in each layer are some of the hyperparameters needed to be optimized in order to have a satisfactory result (3).

##### Random forest

Decision trees are supervised ML algorithms with a flow-chart like structure that work by recursively partitioning the feature space in a top to down manner. Decision trees include internal nodes which indicates a certain feature and the decision rules coming from these nodes, also known as branches. The topmost node (the root node) leads to subsequent nodes and ultimately to the leaf nodes, which hold the final predictions. Random forest (RF) aims to ensemble multiple weak decision trees to create a more robust model. In RF, each tree is trained independently on a bootstrapped sample of the training data. For classification purposes, the final outcome is made based on the class voted by the majority of decision trees (4).

#### A brief introduction of convolutional neural networks

Deep learning (DL) is a broad term applied to ML algorithms that are based on deep NNs, i.e., NNs typically with three or more hidden layers. DL has gained much attention during recent years as it has yielded remarkable results in various applications, such as natural language processing, speech recognition, and computer vision. An important contributor of the high performance of DL-based algorithms in computer vision are CNNs. The idea of CNNs is very similar to the way animal visual cortex processes the visual signals; lower-level neurons capture simple features like edges and corners, while higher-level cells detect more complex patterns, such as shape and texture. Generally, CNNs consist of three types of layers, namely convolutional, pooling, and fully connected layers. Convolutional layers, as the name suggests, work by convolving a set of weights organized in small matrices of different sizes (e.g., 3 × 3, 5 × 5, or 7 × 7) over the input data until all regions are covered. The matrices are also known as filters or kernels and are essentially feature detectors. The convolution operation results in a number of feature maps, each specific for a certain feature of the input data, which are then utilized to be convolved with the filters of the next layer and so on; therefore, simple to higher level features become extracted consecutively through the convolutional network. Pooling layers reduce the spatial dimensionality of the input data and make them invariant to small translation; therefore, an image containing a cat is still labelled as a cat even if the cat location becomes shifted, for example. Pooling layers are mainly composed of max pooling and average pooling layers. In max pooling, the maximum values of each input feature map, corresponding to the most salient features, are selected, whereas in average pooling, all elements of a feature map are averaged. In a typical CNN architecture, several convolutional layers, non-linearity functions like ReLU, and pooling layers are stacked together, and this unit may also become repeated through the network. FC layers are located at the end of a CNN architecture and are similar to a traditional NN. FC part aims to utilize high level representations, generated by convolutional and pooling layers, to predict the class of input data, if a classification problem was dealt with. Training of a CNN involves updating the weights of convolutional and FC layers through the process of backpropagation so that the difference between the actual and predicted class is minimized (3).

#### A brief explanation of the proposed convolutional autoencoder architecture

Briefly, the encoder part was composed of five convolutional blocks and four max pooling layers with a stride of 2 in between for down-sampling. Each convolutional block consists of three successive 2D convolutional layers with a kernel size of 3 × 3 and a same number of channels, progressively increasing from 32 to 64, 128, 256, and finally 512 channels in the first to last encoder block, respectively. In each encoder block, the number of channels doubles in the first 2D convolutional layer, and subsequent convolution operations maintain the same number of channels. Following a final max pooling layer at the end of the encoder path, the bottleneck part of CAE, identical in architecture to the encoder blocks, generates a 4x4x1024 feature map. The feature map is then up-sampled through a transposed 2D convolutional operation with a stride of 2, and it proceeds to the decoder path. In a symmetric way with the encoder path, the decoder compromises five decoder blocks with up-sampling performed between them, again using a transposed 2D convolution operation (stride 2). Decoder blocks 1, 3, and 5 consisted of three consecutive 2D convolutional layers that receive an input map with 512, 128, and 32 number of channels, respectively, and generate maps of a same depth. To prevent losing precise information through up-sampling, the output feature maps obtained from the encoder blocks 2 and 4 are concatenated with the feature maps of the corresponding layers in the decoder path using skip connections. In decoder blocks 2 and 4, a convolution operation first halves the number of channels, and subsequently, two successive convolutional layers, not altering the number of channels, are added. Finally, a 1×1 convolution operation is applied to the output of last decoder block, creating an image of the same size as the input image, i.e., 128×128×1.

#### Figures

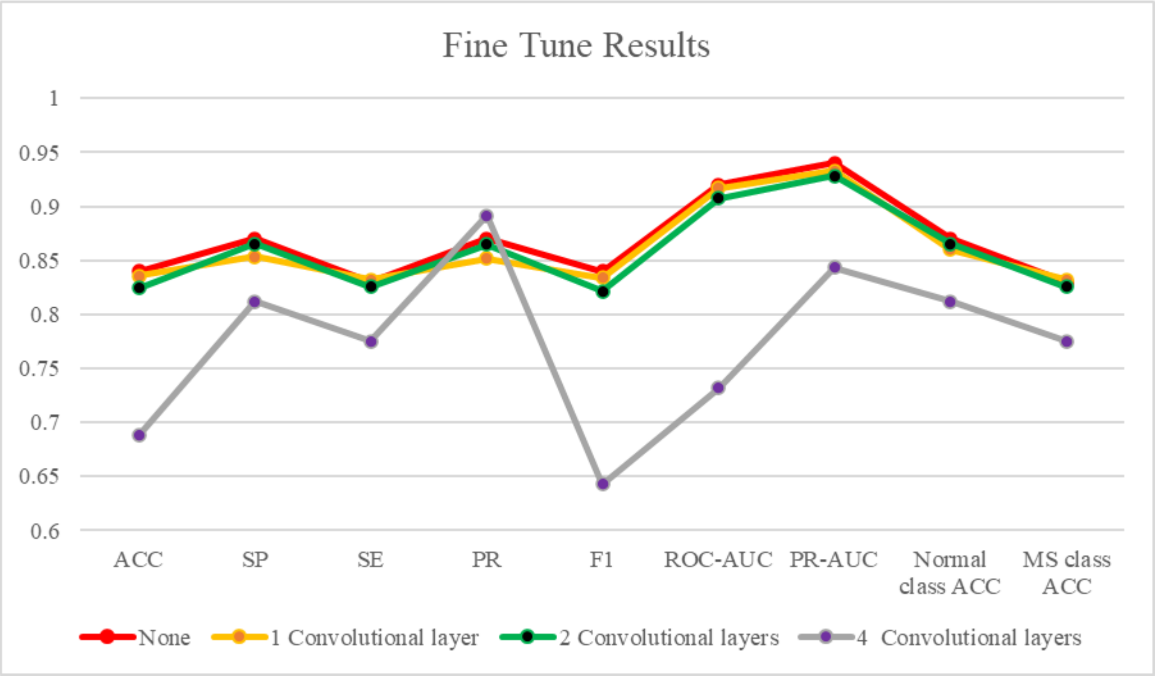


Figure A. performance metrics achieved by fine tuning the best convolutional neural network model, i.e., VGG-19, in which the weights of the first, second, and fourth convolutional layers were unfrozen.

### Tables

Table A. performance metrics of the support vector machine classifier with linear, polynomial, radial basis function (RBF), and sigmoid kernels, applied to the features extracted using the proposed convolutional autoencoder neural network. The optimal hyperparameters are depicted in the last two columns. RBF is the winner.

| Model | Kernel | ACC | SE | SP | PR | F1 | ROC-AUC | PR-AUC | Optimal hyperparameters | |
|---|---|---|---|---|---|---|---|---|---|---|
| **CAE + SVM** | Linear | 0.85 | **0.86** | 0.84 | 0.85 | 0.85 | **0.93** | 0.93 | C | 1 |
| | Polynomial | 0.84 | 0.83 | 0.85 | 0.85 | 0.84 | 0.92 | 0.93 | C | 10 |
| | | | | | | | | | Degree | 3 |
| | RBF | **0.86** | 0.82 | **0.90** | **0.90** | **0.86** | **0.93** | **0.94** | C | 10 |
| | | | | | | | | | Gamma | $10^{-4}$ |
| | sigmoid | 0.71 | 0.61 | 0.83 | 0.79 | 0.71 | 0.78 | 0.84 | C | 1 |

Table B. optimal hyperparameters for RF, MLP), and K-NN classifiers applied to the features extracted using the proposed convolutional autoencoder neural network.

| Model | Hyperparameter | Best value |
|---|---|---|
| **KNN** | number of neighbors | 6 |
| | Weights | Distance |
| | P (power parameter) | 2 |
| | algorithm for nearest neighbor | Auto |
| **RF** | n_estimators (number of trees) | 265 |
| | criterion (the quality of a split) | Entropy |
| | max_depth | 8 |
| | min_sample_split. | 9 |
| | min_sample_leaf. | 5 |
| **MLP** | batch-size | 32 |
| | learning rate | 0.001 |
| | Epoch | 200 |
